# LLIN Evaluation in Uganda Project (LLINEUP) – *Plasmodium* infection prevalence and genotypic markers of insecticide resistance in *Anopheles* vectors from 48 districts of Uganda

**DOI:** 10.1101/2023.07.31.23293323

**Authors:** Amy Lynd, Samuel Gonahasa, Sarah G Staedke, Ambrose Oruni, Catherine Maiteki-Sebuguzi, Penny Hancock, Erin Knight, Grant Dorsey, Jimmy Opigo, Adoke Yeka, Agaba Katureebe, Mary Kyohere, Janet Hemingway, Moses R Kamya, Daniel McDermott, Eric R. Lucas, Martin J Donnelly

**Affiliations:** Liverpool School of Tropical Medicine, Pembroke Place, Liverpool L3 5QA, UK; Infectious Diseases Research Collaboration, 2C Nakasero Hill Road, Kampala, Uganda; Imperial College, Norfolk Place, London, W2 1PG.; University of California, San Francisco, San Francisco, CA 94110 USA; Uganda Ministry of Health, Kampala, Uganda; Makerere University College of Health Sciences Infectious Diseases Research Collaboration, 2C Nakasero Hill Road, Kampala, Uganda

**Keywords:** malaria, long-lasting insecticidal nets (LLINs), piperonyl butoxide (PBO), Uganda, cluster-randomised trial, vector control, insecticide resistance

## Abstract

**Background:** In 2017-2019, we conducted a large-scale, cluster-randomised trial (LLINEUP) to evaluate long-lasting insecticidal nets (LLINs) treated with a pyrethroid insecticide plus the synergist piperonyl butoxide (PBO LLINs), as compared to conventional, pyrethroid-only LLINs across 104 health sub-districts (HSDs) in Uganda. In LLINEUP, and similar trials in Tanzania, PBO LLINs were found to provide greater protection against malaria than conventional LLINs, reducing parasitaemia and vector density. In the LLINEUP trial, cross-sectional entomological surveys were carried out at baseline and then every 6 months for two years. In each survey, ten households per HSD were randomly selected for indoor household entomological collections.

**Results:** Overall, 5395 female Anopheles mosquitoes were collected from 5046 households. The proportion of mosquitoes infected with *Plasmodium falciparum* did not change significantly over time, while infection with non-*falciparum* malaria decreased in *An. gambiae* s.s, but not *An. funestus*. The frequency of genetic markers associated with pyrethroid resistance increased significantly over time, but the rate of change was not different between the two LLIN types. The knock-down resistance (*kdr*) mutation *Vgsc*-995S declined over time as *Vgsc*-995F, the alternative resistance mutation at this codon, increased. *Vgsc*-995F appears to be spreading into Uganda.

**Conclusions:** Distribution of LLINs in Uganda was associated with reductions in parasite prevalence and vector density, but the proportion of infective mosquitoes remained stable, suggesting that the potential for transmission persisted. The increased frequency of markers of pyrethroid resistance indicates that LLIN distribution favoured the evolution of resistance within local vectors and highlights the potential benefits of resistance management strategies.

*Trial registration: This study is registered with ISRCTN, ISRCTN17516395. Registered 14 February 2017,* http://www.isrctn.com/ISRCTN17516395

## Background

Long-lasting insecticidal nets (LLINs) are the principal tool for malaria control in Africa and were the major driver of the decline in malaria mortality between 2000-2015 (Bhatt et al. 2015; World Health Organization 2016). Growing concerns over the rapid spread of resistance to the pyrethroid insecticides with which the nets are treated (Hemingway et al. 2016; Glunt et al. 2018) have been partially assuaged by promising trials of new generation LLINs treated with the synergist piperonyl butoxide (PBO) (Protopopoff et al. 2018; Tiono et al. 2018; Staedke et al. 2020; Maiteki-Sebuguzi et al. 2022), which inhibits the activity of cytochrome P450s, one of the major causes of insecticide resistance in malaria vectors. The LLIN Evaluation in Uganda Project (LLINEUP), was a cluster randomised trial of conventional LLINs (PermaNet 2.0 and Olyset) and PBO-LLINs (PermaNet 3.0 and Olyset Plus). The trial found that parasite prevalence, adjusted for baseline, was 20% lower in children from communities receiving PBO LLINs than in those receiving conventional LLINs, for up to 25 months post-LLIN distribution (parasite prevalence ratio 0·80 [95% CI 0·69–0·93], p=0·0048) (Maiteki-Sebuguzi et al. 2022). Vector abundance, again adjusted for baseline, was 73% lower in PBO communities than in non-PBO communities (Vector density ratio 0·27 [95% CI 0·21–0·36], p<0·0001)(Maiteki-Sebuguzi et al. 2022)

In this paper we report on the frequency of molecular markers of insecticide resistance between the study arms, and over the course of the trial. Insecticide resistance is widespread in malaria vectors in Uganda (Mawejje et al. 2013; Abeku et al. 2017; Okia et al. 2018; Lynd et al. 2019; Njoroge et al. 2022), and has been implicated in the limited epidemiological impact of vector control campaigns (Kigozi et al. 2012; Katureebe et al. 2016; Epstein et al. 2022). The WHO has identified insecticide resistance management (IRM) as a key strategy to prolong the effective lifespan of insecticide-based interventions (World Health Organization 2012). A central tenet of IRM programmes is that use of insecticides within the public health sector selects for resistance and therefore the rational deployment of resistance management tools through rotations, mixtures and mosaics *etc.* can delay the onset of resistance. However, it is unclear whether the application of insecticides for public health is the primary driver of insecticide resistance, or whether use of insecticides for agriculture contributes. Demonstrating the role of the former is important for developing IRM strategies.

The scale and diverse ecologies over which the LLINEUP trial was conducted afforded us the opportunity to test three hypotheses:

> H_1_-the use of pyrethroid insecticides on LLINs selects for an increase in frequency of molecular markers of pyrethroid resistance over the course of the intervention.

A key assumption of IRM strategies is that the use of insecticide synergists, such as PBO, should retard the evolution of resistance. We examine whether there was any evidence for differences in resistance marker frequency changes between intervention arms receiving conventional LLINs and those receiving PBO-LLINs. A priori we proposed the following hypothesis:

> H_2_. The rate of change in frequency in cytochrome P450 resistance markers will differ in *An. gambiae s.s.* populations from clusters which received PBO-LLINs vs conventional LLINs

One of the resistance markers we genotype, the *Cyp6aap-Dup1* haplotype, is more strongly associated with resistance to Class II pyrethroids (including deltamethrin and α-cypermethrin) than to Class I pyrethroids (permethrin) (Njoroge et al. 2022). We therefore propose:

> H_3_. The rate of increase in the *Cyp6aap*-Dup1; ZZB-TE, *Cyp6p4*-236M triple mutant haplotype will be greater in *An. gambiae s.s.* populations from clusters which received conventional deltamethrin LLINs (PermaNet 2.0) relative to clusters which received conventional permethrin LLINs (Olyset).

## Methods

### Household selection, mosquito collection and processing

Full details of the sampling procedures are given in (Lynd et al. 2019; Staedke et al. 2019), Figure 1. In brief, in each round of surveys, 50 households were randomly selected from an enumeration list of households in each of the 104 health sub-districts (HSDs) for the cross-sectional community surveys. Of those 50 households, 10 were randomly selected to take part in the entomology surveys, giving a maximum of 1040 households for entomological surveillance. In the final round of surveys (25 months post net distribution) it was only possible to survey 90 of the 104 HSDs due to restrictions resulting from the COVID-19 pandemic. Mosquitoes were collected using Prokopack aspirators (Vazquez-Prokopec et al. 2009) and DNA extractions were carried out on the head and thorax using Nexttec Biotechnologie DNA extraction plates (Nexttec Biotechnologie GmbH, Hilgertshausen, Germany). *Anopheles gambiae s.l.* and *An. funestus s.l.* mosquitoes were identified to species level by PCR (Koekemoer et al. 2002; Santolamazza et al. 2008). Malaria infections in *An. gambiae s.l.* and *An. funestus s.l.* were detected by a *P. falciparum*, *P. vivax*, *P. ovale* and *P. malariae* Taqman assay (Bass et al. 2008)*. An. gambiae s.s.* females were screened for pyrethroid resistance mutations; *Vgsc*-995F, *Vgsc*-995S, *Vgsc*-1570Y, *Cyp6aap*-Dup1 triple mutant haplotype (consisting of *Cyp6aap*-Dup1 itself, *Cyp6p4*-236M and the transposable element insertion ZZB), *Cyp4J5*-43F and *Coeae1d* following standard protocols (Bass et al. 2007; Jones et al. 2012; Lynd et al. 2018; Weetman et al. 2018; Njoroge et al. 2022). The 2La chromosome inversion karyotype of *An. gambiae s.s.* specimens was assessed by PCR (White et al. 2007).

**Figure. 1:**
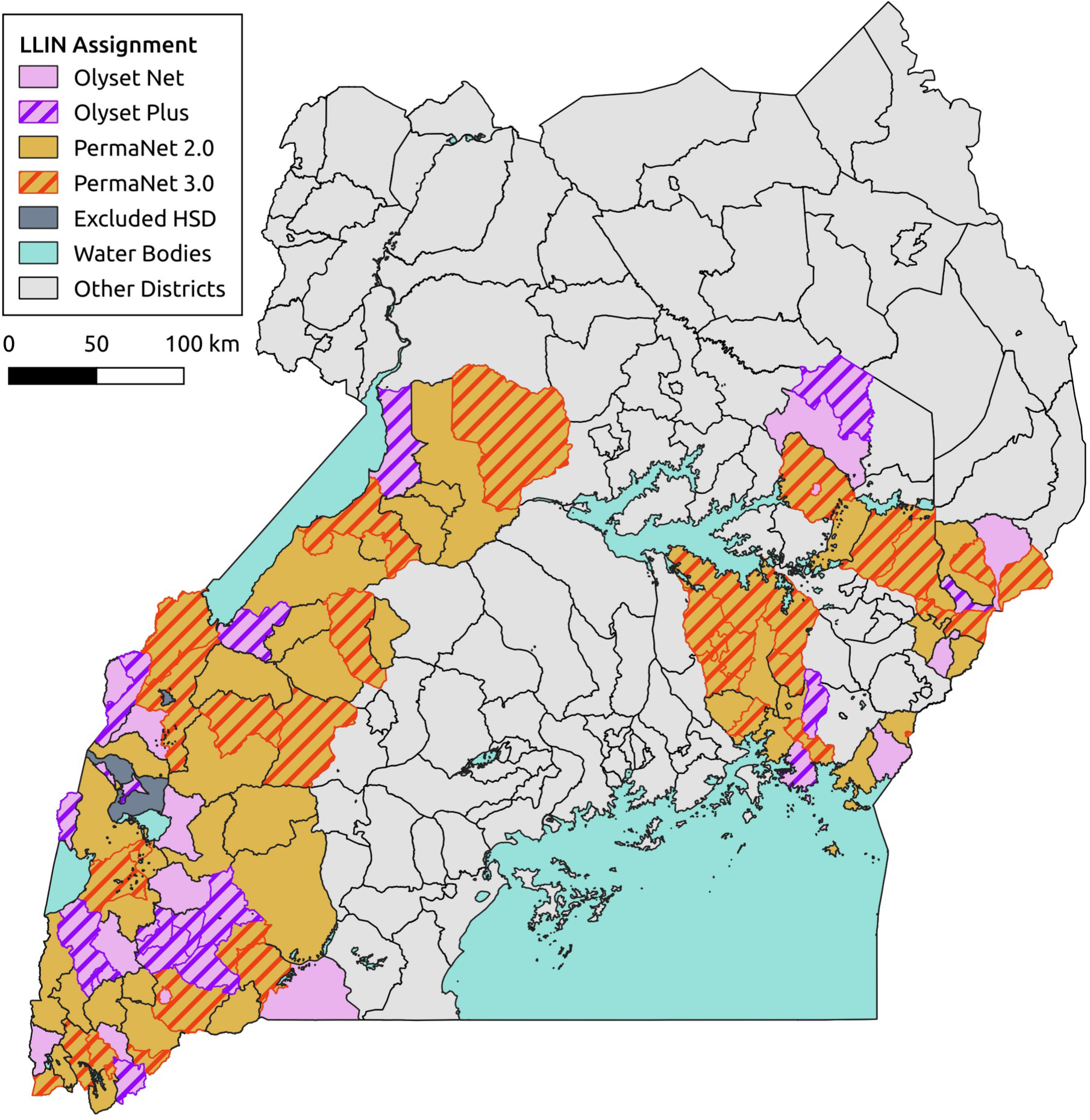
Map of mosquito collection locations within the LLINEUP cluster-randomised control trial. Intervention arm is indicated by colour with the 14 clusters which were omitted during final collection round (25 months post LLIN distribution) indicated with hatched shading.

Data analysis was carried out in R statistical software version 4.1.3 (R Core Team 2015). Analysis of molecular marker frequency data used General Linear Models with logit link function for a binomial dependent variable, implemented in the R package glmmTMB (Brooks et al. 2017). Spatiotemporal variation in marker frequencies was analysed by fitting a Bayesian geostatistical model to the frequencies of each marker observed in the mosquitoes collected from each household in rounds 1-5 (Hancock et al. 2022). The numbers of each allele present in each sample were assumed to follow a binomial distribution, with a mean probability modelled as a spatiotemporal random effect depending on latitude, longitude and round. Spatial autocorrelation was modelled using a Gaussian Markov random field and temporal autocorrelation was an autoregressive model of order 1. Models were fitted using the R-INLA package (Rue et al. 2009; Lindgren et al. 2011).

*Anopheles* household abundance data were reported previously in the main trial papers (Staedke et al. 2020; Maiteki-Sebuguzi et al. 2022) with molecular data from baseline collections published in the baseline entomology paper (Lynd et al. 2019). All data from the 6, 12, 18 and 25 month collection rounds and associated analyses are novel to this manuscript. All collection data and analytical routines are available on GitHub (https://github.com/vigg-lstm/llineup-genotyping).

## Results

Overall, 5395 female Anopheles mosquitoes were collected from 5046 households in the five surveys (Table 1), including 1797 in round 1 (baseline survey), 423 in round 2 (6-months post-LLIN distribution), 755 in round 3 (12-months), 1093 in round 4 (18-months) and 1327 in round 5 (25-months). At baseline, the prevalence of *Plasmodium falciparum* sporozoite infection in *An. gambiae* s.s. was 5.6% (72/1284) and in *An. funestus* was 3.5% (15/432). Other plasmodium species (*P. vivax*, *P. ovale*, and *P. malariae*) were detected less commonly in both *An. gambiae* s.s. (1.2%, 16/1284) and *An. funestus* (1.4%, 6/432) (Lynd et al. 2019). No *Plasmodium* infections were detected in *An. arabiensis* (Supplementary Table 1) and they were excluded from further analysis. The prevalence of *P. falciparum* infection in *Anopheles* mosquitoes did not change significantly over the course of the study. In contrast, the combined prevalence of other *Plasmodium* species decreased significantly in *An. gambiae,* but not *An. funestus* (Figure 2; Table 2). There was no evidence for a significant interaction between collection round and study arm (ie, a difference in the slope of infection prevalence over time between intervention arms) for either *P. falciparum* or other non-falciparum *Plasmodia*.

**Table 1:**
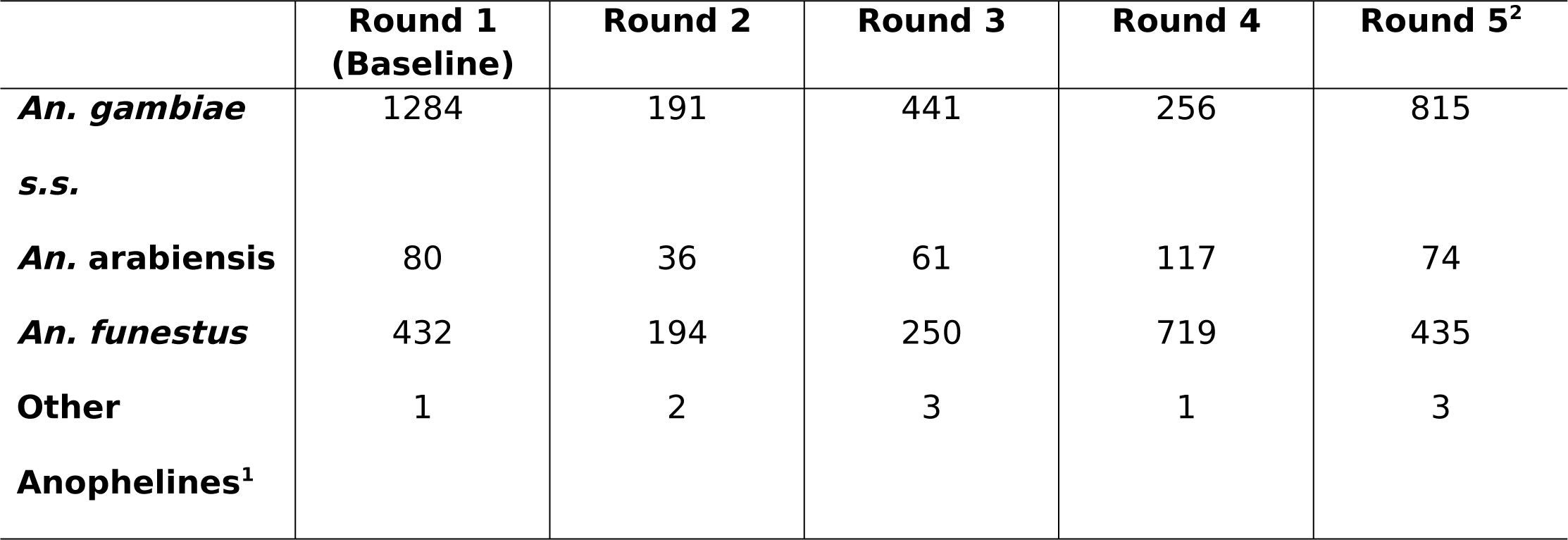
Numbers of female mosquitoes collected from 104 health sub districts across all 5 collection rounds. ^1^ includes *An. coluzzii, An. parensis, An. leesoni, An. rivulorum* and *An. mouchetti.* ^2^Data from only 90 HSDs due to COVID-19 impacts.

**Figure 2.**
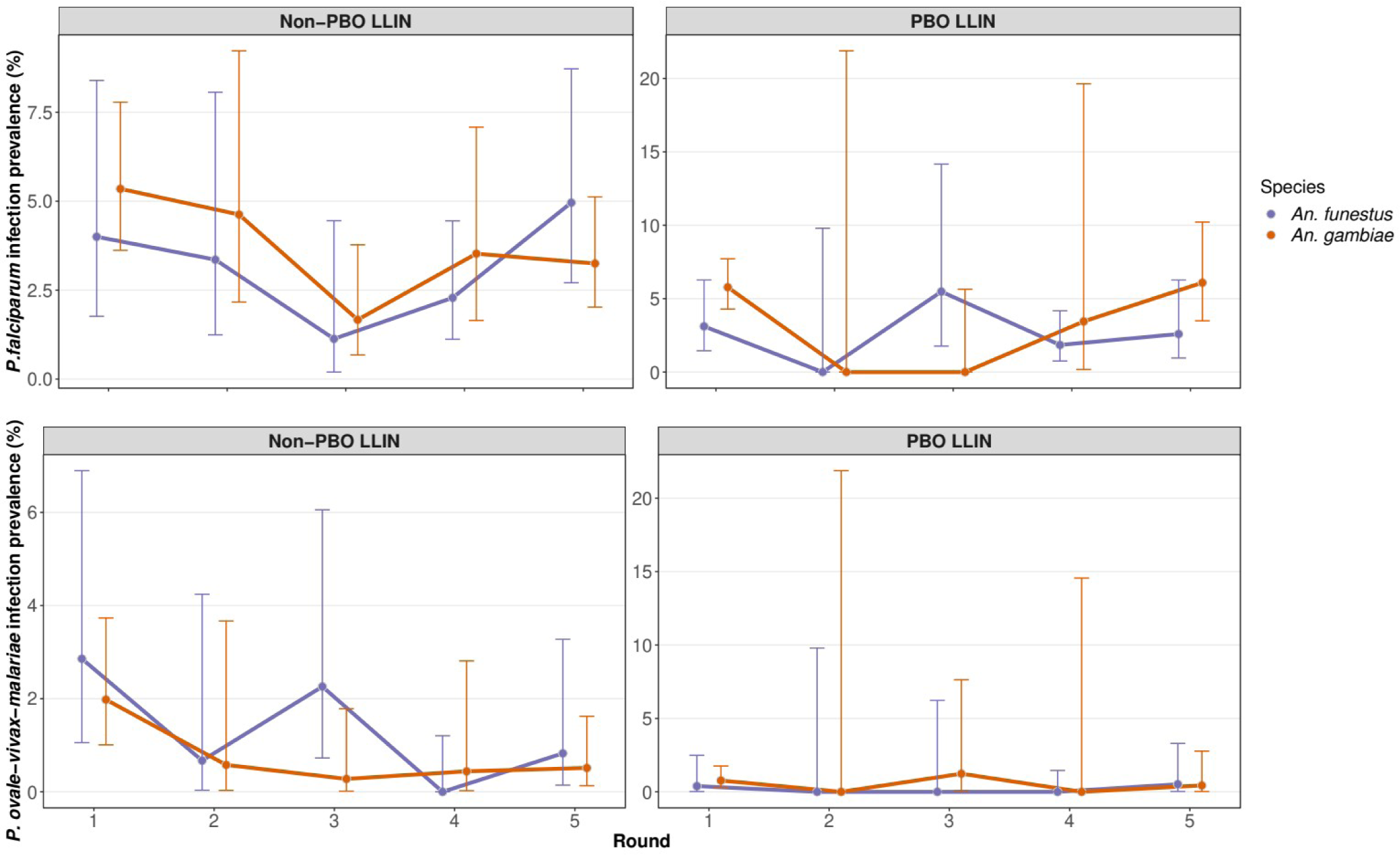
*Plasmodium* infection prevalence in *An. gambiae and An. funestus*. Point prevalence estimate is shown with associated 95%CIs. Data were collected simultaneously but are plotted offset for ease of viewing. Round 1 was the baseline collection with follow up rounds at approximately six-monthly intervals (see text).

**Table 2:** GLMM analysis of *Plasmodium* infection prevalence in the two main vector species. ^1^Model coefficient and significance value. Model 1-Glmm (*P. falciparum* ∼ Round + Arm+ (1|HSD)); Model 2-Glmm (*P. OVM* ∼ Round + Arm+ (1| HSD)). For both models a Round:Arm interaction term was also included but was not found to improve model fit. The “Arm” term distinguishes conventional LLINs and PBO-LLINs but not between deltamethrin and permethrin-treated nets.

|  | **Variable tested** | ***An. gambiae s.s.*<sup>1</sup>** | ***An. funestus*<sup>1</sup>** |
| --- | --- | --- | --- |
| Model 1<br>(Pfal) | Round | -0.107; NS | 0.013; NS |
|  | Arm | 0.259; NS | -0.172; NS |
| Model 2<br>(OVM) | Round | -0.336; p=0.016 | -0.246; NS |
|  | Arm | -0.548; NS | -1.682; p=0.046 |

We screened for insecticide resistance markers only in *An. gambiae* s.s. as it is the primary malaria vector in Uganda, and the species for which the most resistance marker data are available. The resistance associated variant *Vgsc*-1570Y was not found in any specimen and is not discussed further. There was no significant association between any of the genotypic markers screened and infection with *P. falciparum* or the other non-falciparum *Plasmodia* (Supplementary Figure 1).

Collections were conducted over 5 rounds, but mosquito numbers dropped sharply after the LLINs were distributed, as previously reported (Staedke et al. 2020; Maiteki-Sebuguzi et al. 2022), with a notable increase by round 5 (Table 1). Thus, few mosquito samples were available for evaluation in intermediate rounds, particularly in communities that received PBO LLINs. We therefore ran the analysis of genotype frequencies both using data from across all rounds, and focusing on comparisons between baseline and Round 5 (Table 3 and Maiteki-Sebuguzi et al. (2022)).

> H_1_-the use of pyrethroid insecticides on LLINs selects for an increase in frequency of molecular markers of pyrethroid resistance over the course of the intervention.

**Table 3:**
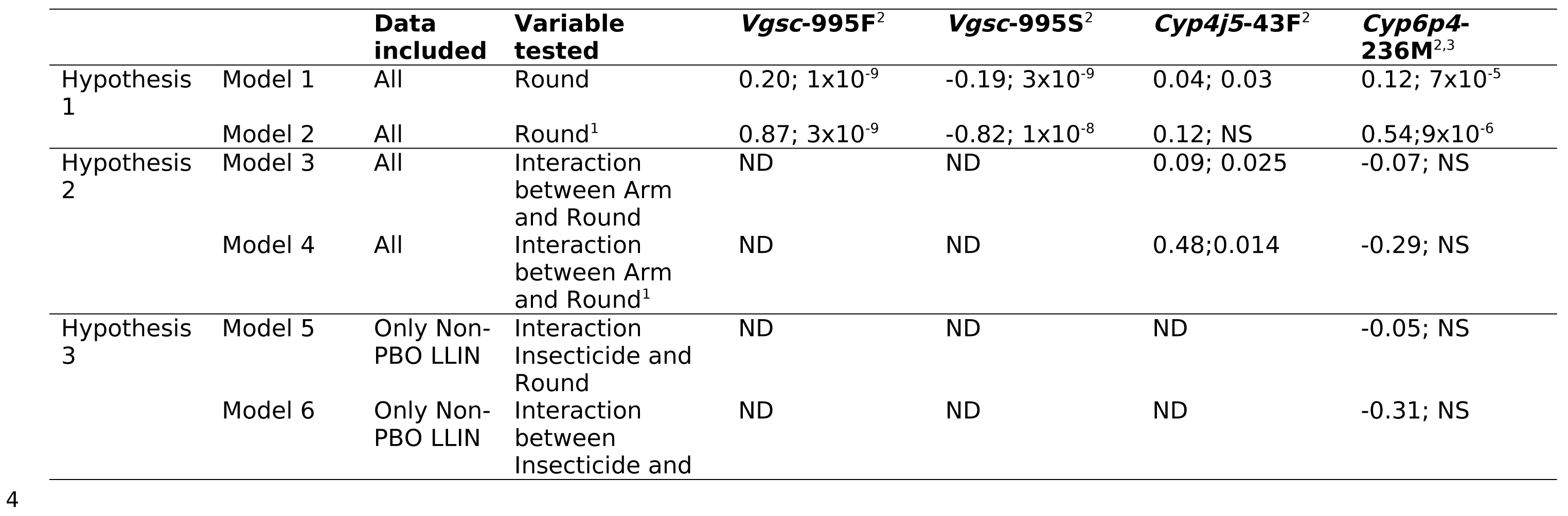
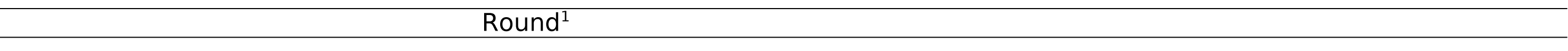
Summary of GLMM analyses of resistance allele frequency changes in *Anopheles gambiae s.s.* ^1^Model compares marker frequency data from baseline and data from 25 months. ^2^Model coefficient and significance value; ^3^Used as the sole marker for the triple mutant haplotype as this marker is in linkage disequilibrium with ZZB-TE and *Cyp6aap*-Dup1. In these collections. *Coeae1d and Chr2La not included as not significant in models 1 and 2 (not relevant for other comparisons) Model 1-Glmm (Marker ∼ Round+ Location+ Arm+ (1|HSD)); Model 2-Glmm (Marker ∼ Round(1vs5)+ Location+ Arm+ (1| HSD)); Model 3-Glmm (Marker ∼ Round+ Location+ Arm+ Round:Arm+ (1|HSD)); Model 4-Glmm (Marker ∼ Round(1vs5)+ Location+ Arm+ Round:Arm+ (1|HSD)); Model 5(Non-PBO-LLINs)-Glmm (Marker ∼ Round+ Location+ Insecticide+ Round:Insecticide+ (1|HSD)); Model 6(Non-PBO-LLINs)-Glmm (Marker ∼ Round(1vs5)+ Location+ Insecticide+ Round:Insecticide+ (1|HSD)). The “Arm” term distinguishes conventional LLINs and PBO-LLINs but not between deltamethrin and permethrin-treated nets. The “Insecticide” term distinguishes between deltamethrin and permethrin-treated nets.

|  |  | **Data included** | **Variable tested** | ***Vgsc*-995F<sup>2</sup>** | ***Vgsc*-995S<sup>2</sup>** | ***Cyp4j5*-43F<sup>2</sup>** | ***Cyp6p4*-236M<sup>2,3</sup>** |
| --- | --- | --- | --- | --- | --- | --- | --- |
| Hypothesis 1 | Model 1 | All | Round | 0.20; 1x10<sup>-9</sup> | -0.19; 3x10<sup>-9</sup> | 0.04; 0.03 | 0.12; 7x10<sup>-5</sup> |
|  | Model 2 | All | Round<sup>1</sup> | 0.87; 3x10<sup>-9</sup> | -0.82; 1x10<sup>-8</sup> | 0.12; NS | 0.54; 9x10<sup>-6</sup> |
| Hypothesis 2 | Model 3 | All | Interaction between Arm and Round | ND | ND | 0.09; 0.025 | -0.07; NS |
|  | Model 4 | All | Interaction between Arm and Round<sup>1</sup> | ND | ND | 0.48; 0.014 | -0.29; NS |
| Hypothesis 3 | Model 5 | Only Non-PBO LLIN | Interaction Insecticide and Round | ND | ND | ND | -0.05; NS |
|  | Model 6 | Only Non-PBO LLIN | Interaction between Insecticide and | ND | ND | ND | -0.31; NS |

Six pyrethroid resistance markers showed a significant change in frequency (*Vgsc*-995S; *Vgsc*-995F; *Cyp4j5*-43F; *Cyp6aap*-Dup1; ZZB-TE, *Cyp6p4*-236M) (Figure 3 and Table 3), either over the course of study follow-up (Table 3 Model 1), or when comparing frequencies at baseline with those from the final collection round (Table 3 Model 2). *Cyp6aap*-Dup1; ZZB-TE, *Cyp6p4*-236M are in near full linkage disequilibrium on a triple-mutant haplotype (Njoroge et al. 2022), so subsequent analyses focused on *Cyp6p4*-236M as representative of this haplotype. No significant changes in frequency for 2La or *Coeae1d* resistance markers were observed. In the 25 months following LLIN distribution, all resistance markers increased significantly in frequency, except for Vgsc-995S which decreased significantly, consistent with the hypothesis that LLINs treated with pyrethroids (a public health intervention) exert selective pressure and drive pyrethoid resistance. The *Vgsc-995S* mutation is in negative linkage with *Vgsc*-995F, with relatively few wild-type alleles in the population (Lynd et al. 2019). Thus, the decrease in *Vgsc*-995S is an expected consequence of the increase in *Vgsc*-995F.

**Figure. 3:**
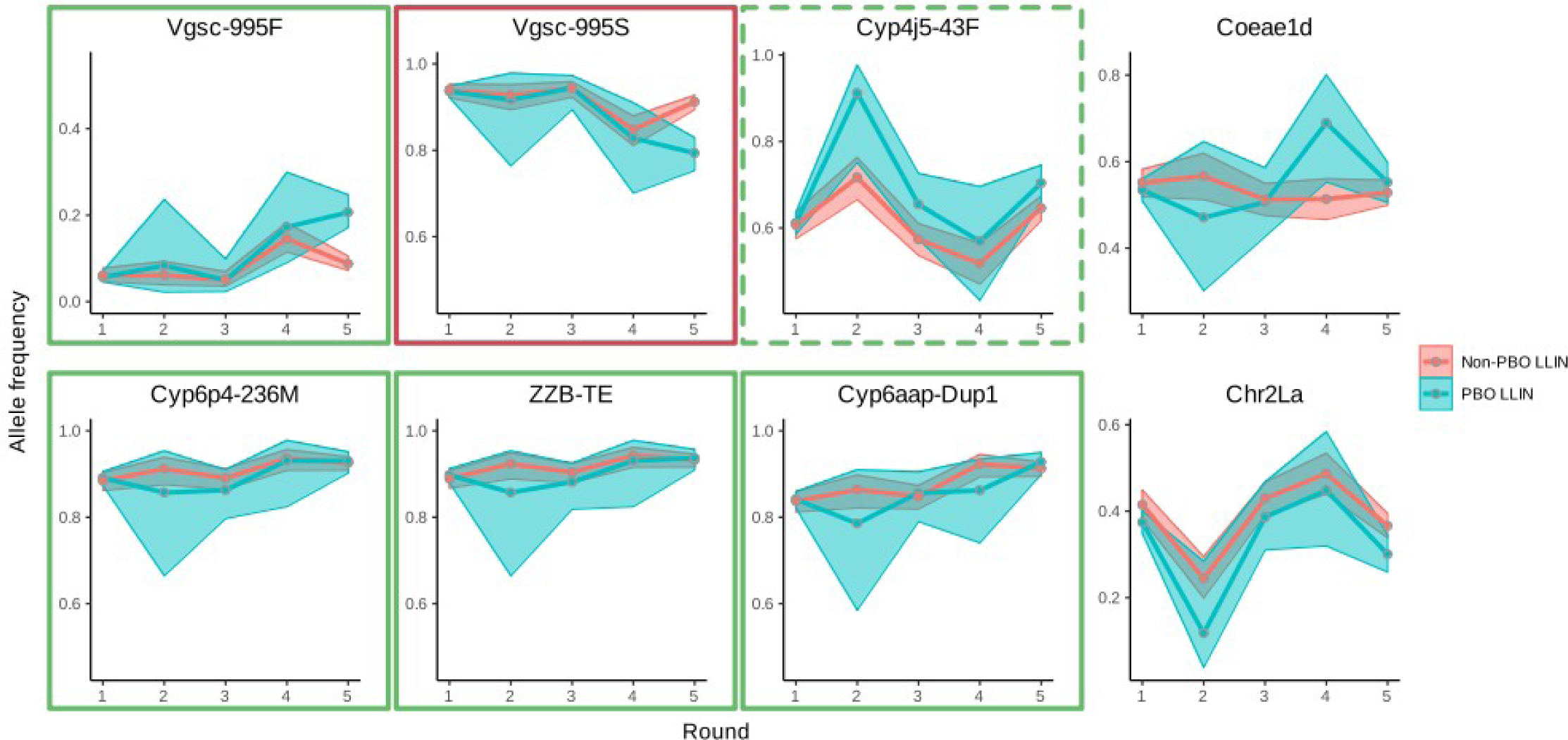
Insecticide resistance marker prevalence *in Anopheles gambiae s.s.* across the baseline and four post LLIN distribution collection rounds. Markers in green external boxes show significant increases (see Table 3) over the course of the trial, red boxes indicate significant decreases (Table 3). Dashed line indicates significant change observed only when GLMM included all five collection points.

Mapping the change in frequency of *Vgsc-*995F suggested that this mutation gradually spread from the North-West of the country over the 2-year study period (Figure 4). In contrast, the increase in frequency of *Cyp4j5*-43F occurred in all areas, and particularly in the Southern regions. At baseline, the *Cyp6p4*-236M (triple-mutant haplotype) was already present at high frequency (>65%) across the study area, and the greatest increases in frequency of this allele were in the North-East, where the baseline frequencies were lowest.

**Figure. 4:**
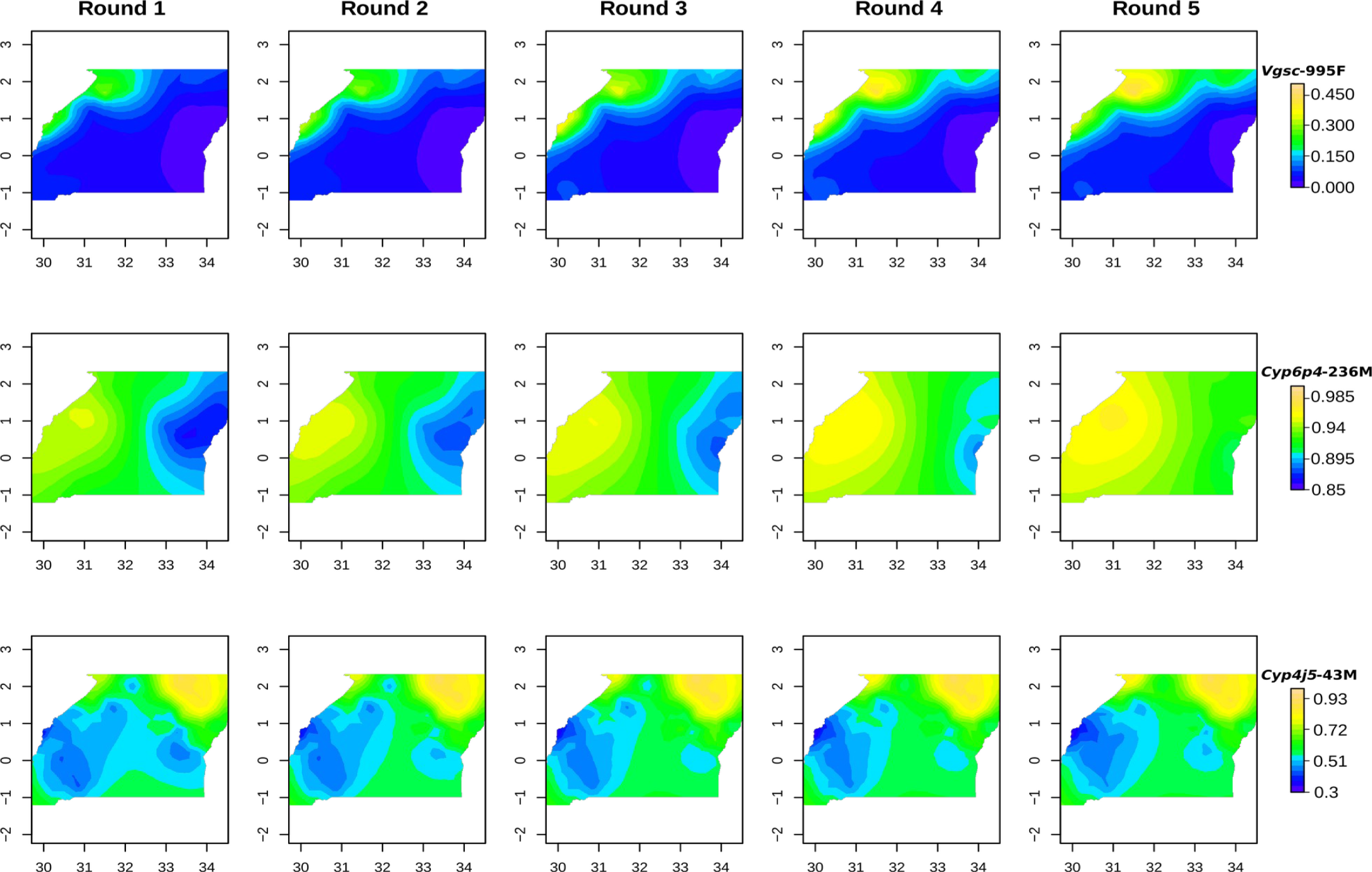
Mapping of mutant allele frequencies over collection rounds for *Vgsc-*995F, *Cyp6p4*-236M and *Cyp4j5*-43F. X and Y axis tickmarks show longitude and latitude respectively. Note colour scales do not carry over across rows.

> H_2_. The rate of change in frequency of cytochrome P450 resistance markers will differ in *An. gambiae s.s.* populations from clusters which received PBO-LLINs vs conventional LLINs

*Cyp4j5*-43F and *Cyp6p4*-236M (triple-mutant haplotype) showed contrasting trends in frequency (Fig. 3). For *Cyp4j5-*43F, the interaction between collection round and LLIN arm was significant, both over the course of the study (Table 3 Model 3) and when comparing pre-LLIN distribution frequencies with the final collection round (Table 3 Model 4). The direction of the interaction indicates that the rate of increase in this P450-mediated resistance mechanism is, perhaps counter-intuitively, higher in clusters which received PBO-LLINs. Conversely there was no evidence for a significant interaction between round and intervention arm for the triple haplotype marker, *Cyp6p4*-236M.

> H_3_. The rate of increase in the triple mutant haplotype will be greater in *An. gambiae s.s.* populations from clusters which receive deltamethrin LLINs (PermaNet 2.0) relative to clusters which receive permethrin LLINs (Olyset).

There was no evidence that rates of change in frequency of the triple-mutant haplotype were different between clusters receiving conventional LLINs treated with either permethrin or deltamethrin (Table 3 models 5 and 6).

## Discussion

To investigate the impact of LLINs on the emergence and spread of insecticide resistance in Uganda, we evaluated *Anopheles* mosquitoes collected from 48 districts over the 2-year follow-up period of the LLINEUP trial. We found that although parasite prevalence and vector density decreased in both study arms following the distribution of LLINs (Maiteki-Sebuguzi et al. 2022), the rate of infection with *Plasmodium falciparum* in sampled mosquitoes did not change significantly over the study, while rates of infection with other *Plasmodium* species decreased significantly in *An. gambiae,* but not *An. funestus*. Since the number of mosquitoes collected following LLIN distribution was low, it is possible that the lack of significance is a result of low statistical power. However, in the case of *An. funestus,* there was no suggestion of an overall decrease in infection over time (the model coefficient was positive). The reduction in parasite prevalence (Maiteki-Sebuguzi et al. 2022) and mosquito numbers does not appear to have markedly reduced sporozoite rates in the *Anopheles* vectors analysed. In the LLINEUP trial, the reduction in parasite prevalence was observed in children aged 2-10 years. It is possible that parasitaemia in older children and adults persisted, thus providing opportunities for mosquitoes to take infected blood meals from this reservoir of older residents. Increased mosquito mortality, inferred from reduced vector collections, is expected to result in a younger mosquito population, leading to a smaller proportion of mosquitoes living long enough to become infective. However, increased adult mortality does not necessarily result in a younger age distribution if it causes a population decline (Abrams 1993), and mosquito infection rate may therefore be unaffected.

We found no evidence of association between resistance marker genotype and infection status. These results contrast to those from a previous study which detected an association between *Vgsc*-995S genotype and infection, consistent with the hypothesis that mosquitoes carrying resistant alleles had increased longevity and were therefore more likely to survive the parasite extrinsic incubation period(Kabula et al. 2016). One difference that could explain these results is that overall *Vgsc*-995 and *Cyp6p4*-236M mutant frequencies were high in our study, with very few wild-type alleles found in the population. There may therefore have been too few fully susceptible individuals to detect an effect of resistance on *Plasmodium* infection.

There was clear evidence of increases in genotypic markers of pyrethroid resistance over the study. Although it not possible to randomise resistance between trial arms, it is most likely that the increases in resistance variants were associated with the distribution of LLINs as the mosquitoes were collected from HSDs representing ≈40% of the surface area of Uganda, encompassing marked differences in ecology, altitude, socio-economic status of communities etc (Maiteki-Sebuguzi et al. 2022). Previous work to correlate LLIN distributions with changes in resistance have yielded contrasting results, arguably due to their reliance upon inherently noisy resistance phenotyping approaches (Implications of Insecticide Resistance Consortium 2018). Our use of genotypic markers provides a metric that can be accurately quantified, reducing the noise in the statistical analysis. The significant increase in *Vgsc*-995F frequency at the expense of the alternative *Vgsc-*995S allele suggests that the former mutation is gradually replacing the latter, and mapping of allele frequencies indicates that this replacement is centred in the North-West of the country. *Vgsc*-995F is a predominantly West- and Central-African allele, and thus its higher frequency in the North-Western part of Uganda is consistent with a gradual spread eastwards. *The Vgsc-*995F was first observed in *An. gambiae s.s.* from the region in 2012 (Ochomo et al. 2015). At this time the *Vgsc*-995S variant was near fixation (Mathias et al. 2011) and analyses suggested that the *Vgsc*-995S mutation was older (Lynd et al. 2010) and more strongly associated with DDT rather than pyrethroid resistance (Reimer et al. 2005; Donnelly et al. 2016). The increase in frequency of the *Vgsc-*995F mutation provides additional support to our contention that LLINs are a major driver of pyrethroid resistance.

PBO LLINs have been promoted as an intervention that can overcome, at least in part, cytochrome P450-mediated resistance. A priori we would have predicted that mosquito populations sampled from communities that received PBO LLINs would show a decrease in P450-resistance markers relative to conventional LLINs. However whilst there were contrasting patterns observed between different P450 marker systems, the only evidence of a significant change was a relative increase in frequency of the *Cyp4j5*-43F marker. It may be that incomplete suppression of cytochrome P450s does not remove the selective pressure to upregulate this form of resistance, but rather escalates an arms race in which mosquitoes further upregulate cytochrome P450-mediated resistance to overcome the suppressive effects of PBO.

## Conclusions

The large-scale deployment of LLINs in Uganda in 2017-2019 was associated with an increase in the frequency of genotypic markers of pyrethroid resistance, but not in the frequency of *P. falciparum* infection in mosquitoes. The reduction in malaria prevalence resulting from the LLIN distribution campaign was therefore likely the result of decreased mosquito numbers, rather than fewer infective mosquitoes. The increase in resistance allele frequency suggests that public health interventions, such as LLINs, can apply selective pressure which drives the evolution of insecticide resistance, supporting the need for resistance management strategies.

## Declarations

### Ethics approval and consent to participate

The study was approved by the Ugandan National Council for Science and Technology (UNCST Ref HS 2176), Makerere University School of Medicine Research & Ethics Committee (SOMREC 2016-133), London School of Hygiene & Tropical Medicine Ethics Committee (LSHTM Ref 12019), and the Liverpool School of Tropical Medicine (Ref 16-072), which sponsored the study. Written informed consent to participate in the study was obtained by the head of household (or their designate) for all participating households.

### Consent for publication

Not applicable

### Availability of data and material

The datasets reported herein are publicly available in the GitHub repository https://github.com/vigg-lstm/llineup-genotyping, as are the scripts used in the analysis of these data.

## Data Availability

All data are available fomr the Github repository cited withing the article

https://github.com/vigg-lstm/llineup-genotyping

## Author’s contributions

SGS, MRK, GD, and MJD conceived of the study, with input from JO, AY, and JH. SGS, GD, and MJD developed the procedures and drafted the protocol with MRK and JH. AL led the data collection in the field, with oversight from SG, and support from CMS, JO, MRK, AK, MK and SGS. AL and AO performed all laboratory analyses. AL, ERL, DMcD, PH, EK and MJD managed the data and led the data analysis. AL, ERL and MJD interpreted the data and drafted the manuscript, with input from SGS. All authors reviewed the manuscript and gave permission for publication. MJD, the corresponding author, had full access to all the data in the study and had final responsibility for the decision to submit for publication.

All authors read and approved the final version of the manuscript.

## Acknowledgements

The authors would like to thank all the staff and administration at IDRC for their considerable efforts in facilitating this project. In particular Martin Mugote, Violet Tuhaise, Jackson Asiimwe and Kilama Maxwell Kilama for their supervision of field activities; Peter Mutungi, and Simon Peter Kigozi and Geoff Lavoy for their considerable assistance in data management; Lillian Taaka, Christine Nabirye and Nicholas Wendo for assistance in logistics and procurement; Susan Nayiga, Isiko Joseph, Erias Muyanda and Henry Opolot for coordination and support. We are extremely grateful to the entomology field workers for collecting and identifying mosquitoes, and to the community survey teams and team leaders who facilitated their work. We would also like to acknowledge and thank the members of the Uganda National Malaria Control Program and the Liverpool School of Tropical Medicine for logistical and other support provided as we carried out these surveys and the district health, administrative, and political leadership teams for all their support and guidance during community entry in the 48 districts of the study area. We thank Dr David Weetman for comments on an earlier version of the manuscript.

---

**Supplementary table 1:**
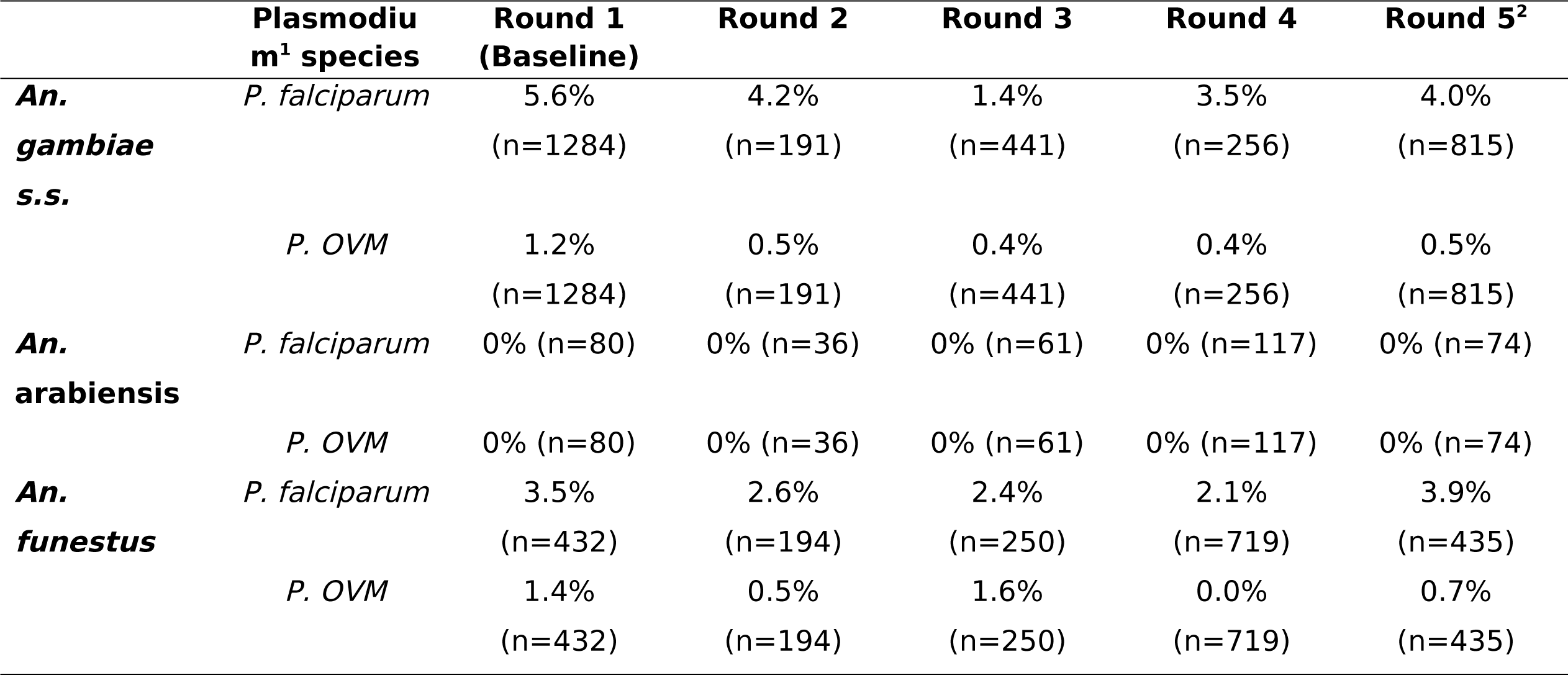
Prevalence of *Plasmodium* positive mosquitoes collected from 104 health sub districts across all 5 collection rounds. ^1^ Data for *Plasmodium falciparum* and for *P. ovale*, *P. vivax* and *P. malariae* combined. ^2^Data from only 90 HSDs due to COVID-19 impacts.

**Supplementary figure 1:**
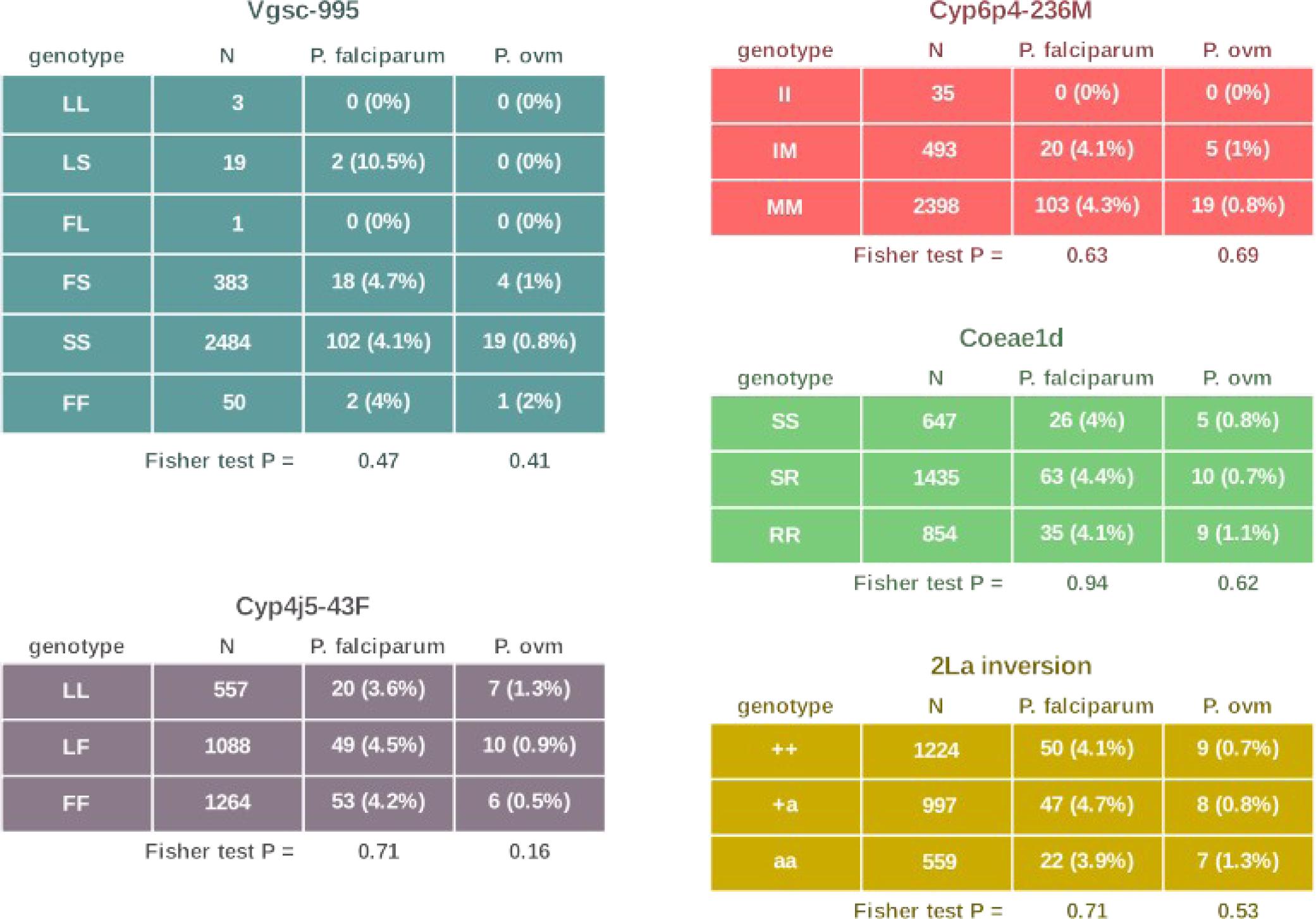
Infection prevalence of *P. falciparum* and *P.ovale + vivax + malariae (ovm)* in *An. gambiae* stratified by resistance associated marker genotype. There was no significant association between infection with either parasite grouping and genotype.

